# Gut Microbiota in Elderly Japanese Patients Undergoing Long-Term tube feeding

**DOI:** 10.1101/2025.08.03.25332055

**Authors:** Yosuke Maruyama, Miyuri Sasaki, Yasuko Ohta, Yukiko fujikawa, Hiroki Tanabe, Yoshiko Hasebe

## Abstract

Tube feeding (TF) is a common nutritional intervention in elderly patients. However, TF may change the composition and diversity of the gut microbiota, resulting in gastrointestinal symptoms such as diarrhea and constipation. This study investigated the impact of TF on the gut microbiota of Japanese individuals aged 60 years or older who had received TF for at least six months. Using 16S rRNA gene sequencing, the gut microbiota profiles of long-term TF (LTF) patients (LTFP) were compared with those of healthy controls (CON). The LTFP group exhibited a significant reduction in bacterial richness and diversity compared with the CON group. Taxonomic analysis revealed a marked decrease in the phylum *Firmicutes* (*p* = 0.0424), families such as *Ruminococcaceae* (*p* < 0.0001), *Prevotellaceae* (*p* = 0.0029), and *Veillonellaceae* (*p* = 0.0017), and genera including *Roseburia* (*p* = 0.0017) and *Subdoligranulum* (*p* = 0.0001) in LTFP group. Functional prediction analysis showed an enrichment of catabolic pathways in LTFP group, particularly of those related to nucleotide degradation and fermentation, whereas the CON group exhibited enriched biosynthetic and maintenance pathways. These findings suggest that LTF may lead to a dysbiotic gut environment, characterized by reduced short-chain fatty acid production and altered metabolic potential. Monitoring microbial changes during TF may be useful in guiding nutritional management strategies, including the use of prebiotics and probiotics, to support gut health and quality of life in elderly patients.

## Introduction

Recently, there has been an increase in the aging population in countries such as Japan, Italy, and Germany. More than ever, elderly people in these countries have physical limitations or dementia that curtail their ability to eat independently. The percentage of the elderly population (aged ≥ 65 years) in these countries was 26% in 2015, and it is expected to exceed 30% in 2025. In Japan, it may reach 39.9% before 2060 (BUREAU, Statistics 2015; Arai et al., 2015). Elderly people eat much less and much less frequently than younger people. During acute and chronic illnesses, decreased food intake is accompanied by energy deficiency and general malnutrition. Malnutrition accounts for 40% of hospitalizations in the elderly. A study by Allison et al. (2000) reported that energy consumption in elderly patients is less than 70% of the recommended intake. However, some elderly patients with reduced feeding function regulate their nutritional status via tube feeding (TF). TF is a common method for providing adequate nutrition in situations where normal oral nutrition is insufficient (e.g., people with significant dysphagia, aspiration, or severe metabolic conditions), and has been used for elderly individuals with advanced dementia, difficulty swallowing, and low nutritional intake (Dharmarajan et al., 2004). The introduction of TF is beneficial for patients with refractory malnutrition, resulting in a significant increase in BMI (Charles et al., 2018).

However, TF has certain limitations. For example, long-term TF (LTF) has been reported to decrease the diversity of gut microbiota, diminishing healthy microbial communities and increasing potentially pathogenic bacteria, accompanied by a reduction in anaerobic bacteria and an increase in aerobic bacteria (Schneider et al., 2000, Schneider et al., 2010). Additionally, gastrointestinal disorders such as constipation and diarrhea have been reported among LTF patients (LTFP) (Cataldi-Betcher et al., 1983; Bittencourt et al., 2012). Gut microbiota is also known to vary widely among racial/ethnic groups and geographical regions, with Central and South Asian populations showing microbiome profiles that differ significantly from those of people from Europe and North America (Abdill et al., 2025).

According to Nishijima et al. (2016), the gut microbiota of the Japanese population differs considerably from that of other populations. It is characterized by a high abundance of bacteria from the phylum *Actinobacteria*, particularly from the genus *Bifidobacterium*, and a predominance of active carbohydrate metabolism pathways,along with a reduction in pathways related to replication, repair, and cell motility. Differences associated with age, sex, and fecal conditions have also been reported (Takagi et al., 2019).

Although several studies have analyzed the gut microbiota of patients receiving TF in diverse countries, such as the United States, China, and France, and across various age groups and disease conditions, to the best of our knowledge, no study has reported the gut microbiota of elderly Japanese individuals undergoing LTF.

Based on previous reports, we hypothesized that LTF in elderly Japanese individuals causes a decrease in diversity and other changes in the bacterial microbiota. To test this hypothesis, we evaluated and compared the gut microbiota of elderly Japanese LTFP and healthy control (CON) individuals through 16S rRNA gene sequencing of their fecal specimens. Our results clarify the effects of LTF on the gut microbiota of elderly Japanese individuals and contribute to LTF optimization.

## Methods

### Participants and Experimental Design

This study was conducted in compliance with the Declaration of Helsinki and Ethical Guidelines for Medical and Health Research. This study was approved by the ethics committees of Nayoro City University (application number 18-035). The characteristics of the participants were obtained from their nursing records. For the CON group, age-matched acquaintances of the researchers were recruited. Informed consent was obtained from all participants.

### Fecal sample collection

A stool sampling kit (TechnoSuruga Lab Co., Ltd., Shizuoka, Japan FS-0006) was used to collect fecal samples from elderly LTFP admitted to Shibetsu City Hospital, Shibetsu city (44.179°N, 142.405°E), Hokkaido, Japan, from November to December 2018. Control fecal samples were simultaneously collected from healthy elderly people in Sapporo city (43.062°N, 141.354°E), Hokkaido, Japan (Fig. 1). All samples were stored in sampling buffer at room temperature until DNA extraction.

**Figure 1.**
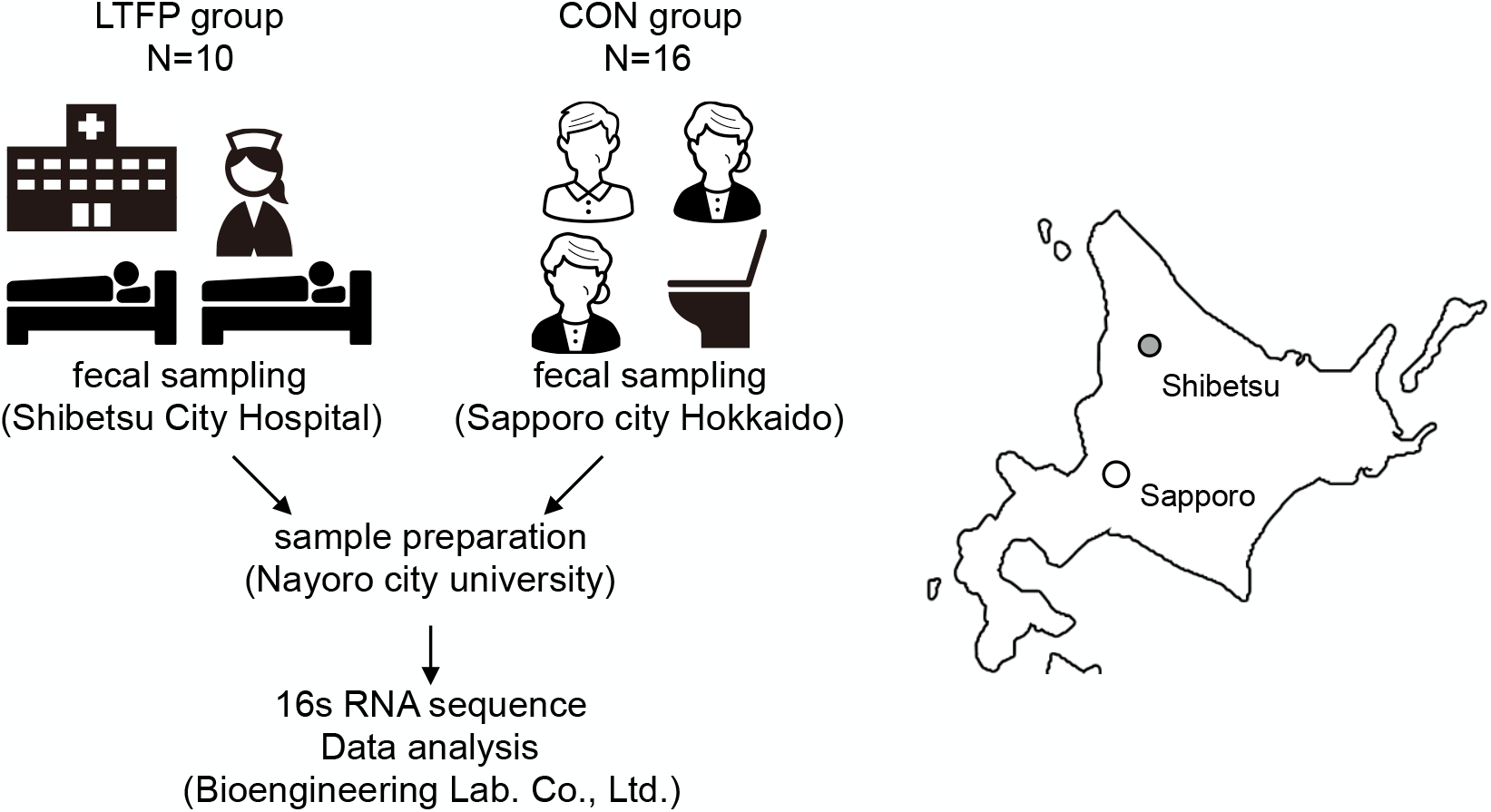
Experimental procedure. Fecal samples from patients receiving LTF were collected at Shibetsu City Hospital, while CON samples were obtained from healthy residents of Sapporo City, Hokkaido, Japan.

### DNA extraction

DNA was extracted using a QIAamp DNA stool mini kit (Qiagen 19590), according to the manufacturer’s protocol. The extracted DNA was quantified with a Quantus™ Fluorometer (Promega E6150) using a QuantiFluor® ONE dsDNA system (Promega E4871). DNA integrity was assessed by 1% agarose gel electrophoresis. Extracted DNA was stored at −80°C until use.

### 16S rRNA gene sequencing

PCR amplification was performed using primers 515F (5^′^-ACACTCTTTCCC TACACGACGCTCTTCCGATCT-NNNNN-GTGCCAGCMGCCGCGGTAA-3^′^) and 806R (5^′^-GTGACTGGAGTTCAGACGTGTGCTCTTCCGATCT-NNNNN-GGACTACNVGGGTWTCTAA-3^′^), targeting the V4 region of the 16S rRNA genes (Caporaso et al., 2011). The reaction mixture contained 0.5 U of KAPA HiFi DNA polymerase (Kapa Biosystems KK2601). PCR amplification was performed according to the following protocol: 94°C for 2 min, followed by 25 cycles at 95°C for 30 s, 55°C for 20 s, and 72°C for 20 s, and a final elongation at 72°C for 5 min. Library construction and sequencing were performed at Bioengineering Lab Co., Ltd. (Atsugi, Japan), and amplicon sequencing was carried out with paired-end tags (2 × 300 bp) on an Illumina MiSeq platform (San Diego, CA, USA). Sequencing data were analyzed using QIIME2 version 2019.0115. Briefly, using Dada2, primers were trimmed, and data were filtered and truncated to allow a 20 bp overlap between forward and reverse reads. Possible chimeric sequences were identified and removed before amplicon sequence variants were assigned using MiDAS database (version 2.1.3).

### Statistical analyses

Results were expressed as the mean ± standard deviation or relative value. Statistical significance was set at p < 0.05. Chao1, Observed species, PD whole tree, and Shannon phylogenetic indices were further analyzed for determining statistical differences using the Wilcoxon rank-sum test. The Wilcoxon test and Kruskal-Wallis rank-sum test were used to identify significant differences between taxa at p < 0.05. Based on 16S rRNA data, microbial functions were inferred and categorized through comparisons using the MetaCyc database and phylogenetic investigation of communities by reconstruction of unobserved states (PICRUSt) along with STAMP (Langille et al., 2013).

## Results

### Sequencing data, and alpha and beta diversity determination

The characteristics of the members of the two groups included in this study, the CON (n = 16) and LTF (n = 10) groups, are summarized in Table 1. 16S rRNA gene sequencing was performed to analyze the gut microbiota of the two groups. In total, we obtained 3,391,683 raw reads from the 26 CON and LTFP samples. The sequencing depth, Q20/Q30 quality scores, and alpha diversity indices (e.g., observed OTUs and Chao1 estimator) for each subject are summarized in Supplementary Table 1. The number of reads per sample ranged from 90,524 to 154,729, whereas the median number of non-chimeric reads was 77,836 (ranging between 53,689 to 105,389). In addition, the Q20 and Q30 scores ranged from 88.3 to 93.4 and 81.3 to 86.9, respectively (Supplementary Table 1). The median number of OTUs was 454, whose values ranged from 290 to 759 (Supplementary Table 1). Alpha diversity analyses, including the observed species (p = 0.081) and the Chao1 (p = 0.549), PD whole tree (p = 0.016), and Shannon (p = 0.097) indices, showed that the species in the LTF and CON groups were significantly different (Fig. 2A).

**Table 1.**
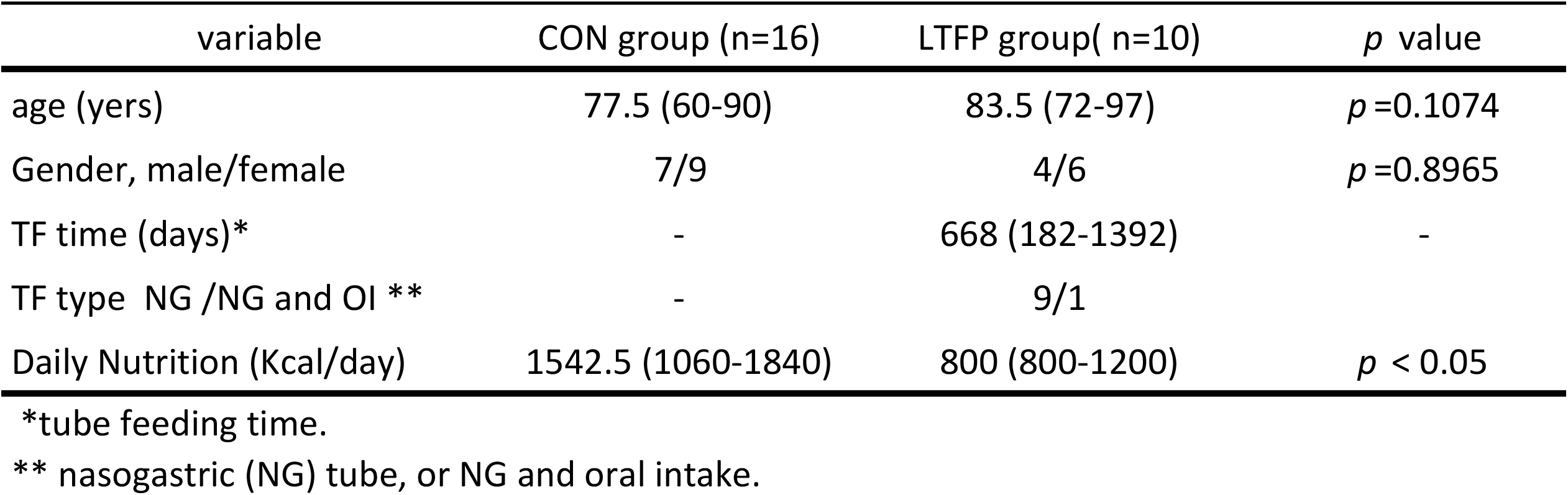
Characteristics of subjects in the CONG and LTFP.

| variable | CON group (n=16) | LTFP group( n=10) | *p* value |
| --- | --- | --- | --- |
| age (yers) | 77.5 (60-90) | 83.5 (72-97) | *p* =0.1074 |
| Gender, male/female | 7/9 | 4/6 | *p* =0.8965 |
| TF time (days)* | - | 668 (182-1392) | - |
| TF type NG /NG and OI ** | - | 9/1 |  |
| Daily Nutrition (Kcal/day) | 1542.5 (1060-1840) | 800 (800-1200) | *p* < 0.05 |

\*tube feeding time.
\*\* nasogastric (NG) tube, or NG and oral intake.

**Figure 2.**
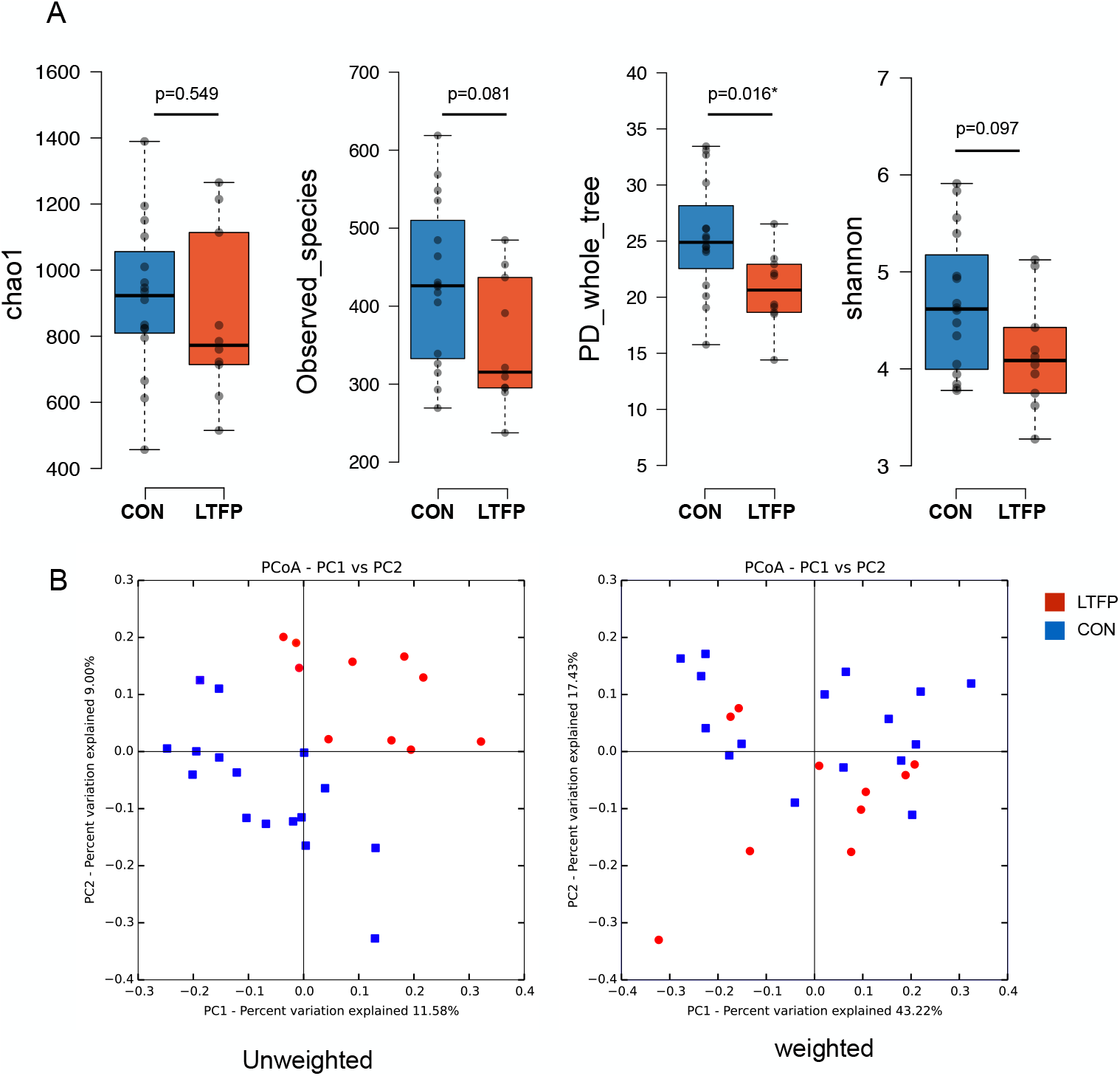
Alpha and Beta diversity metrics analysis. (A) chao1 index, observed species, PD whole tree and Shannon index. (B) Principal component score plots based on unweighted and weighted.

Further, to evaluate the overall differences in beta diversity between the gut microbiota of the groups, we applied Principal Coordinate Analysis (PCoA), based on unweighted and weighted UniFrac distances for the sample set. In the unweighted PCoA, PC1 and PC2 explained 11.85% and 9.0% of the total variation, respectively; whereas, in the weighted PCoA, PC1 and PC2 explained 43.22% and 17.43% of the total variation, respectively (Fig. 2B). PCoA plots suggest distinct clustering patterns between the control (CON) and test groups, indicating potential functional differences in gut microbiota composition between the two groups.

### Relative abundance of gut microbiota in LTFP

The microbial composition of the fecal samples from the CON and LTFP groups was visualized using a stacked bar chart that showed that the relative bacterial abundance at the phylum, class, order, family, and genus levels differed between the two groups (Fig. 3). The mean relative abundances of the major bacterial phyla, classes, orders, families, and genera in each group are listed in Table 2. Detailed values for all samples are provided in Supplementary Table 2. At the phylum level, *Bacteroidetes* accounted for the major bulk of the gut microbiota (47.3% and 50.4% in the CON and LTFP groups, respectively), followed by *Firmicutes* (34.8% and 22.4% in the CON and LTFP groups, respectively), *Proteobacteria* (12.36% and 15.1% in the CON and LTFP groups, respectively), and *Verrucomicrobia* (0.15% and 6.39% in the CON and LTFP groups, respectively) (Table 2).

**Table 2.**
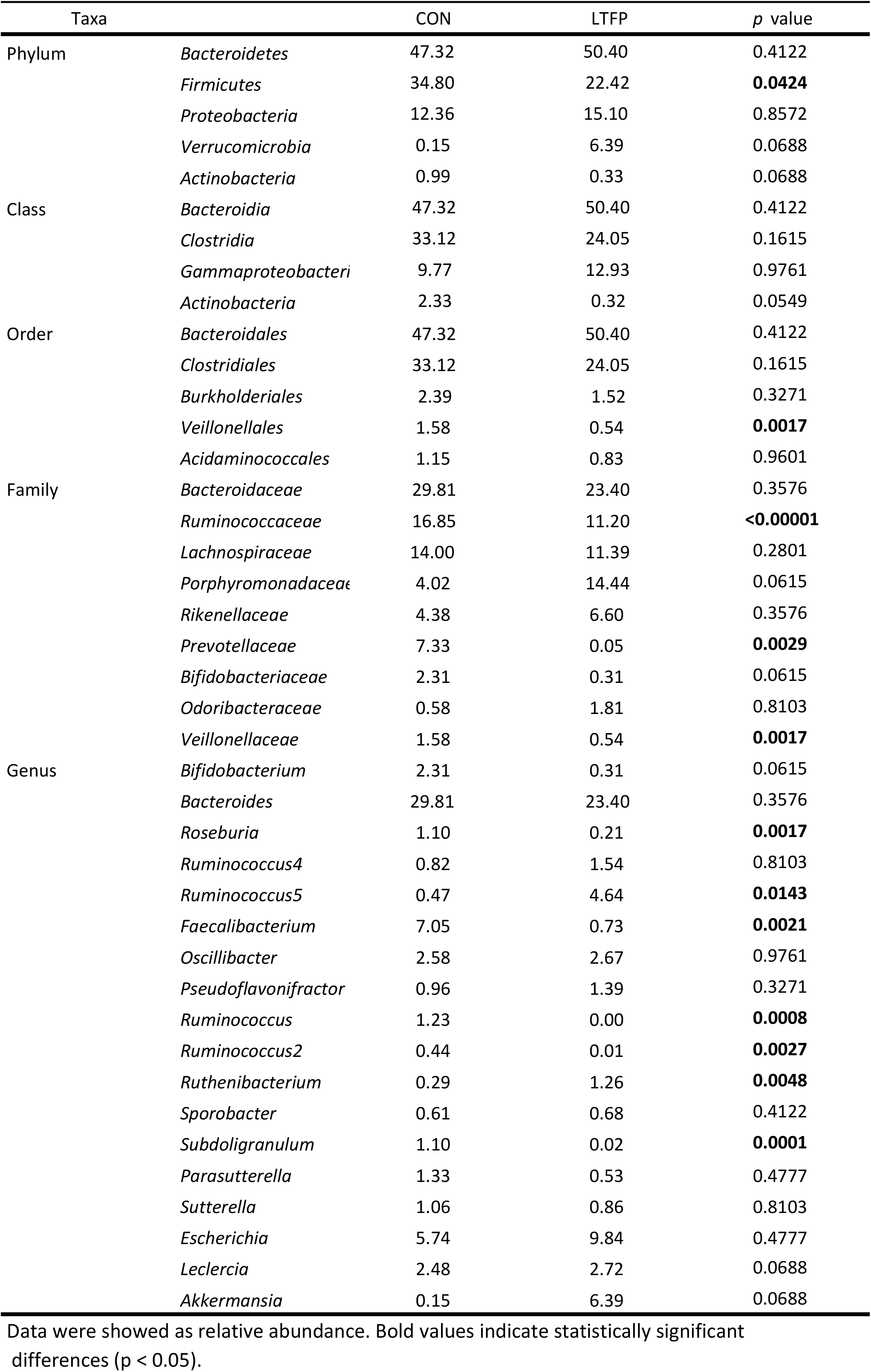
Comparison of relative abundance of fecal microbiota CON and LTFP groups.

**Figure 3.**
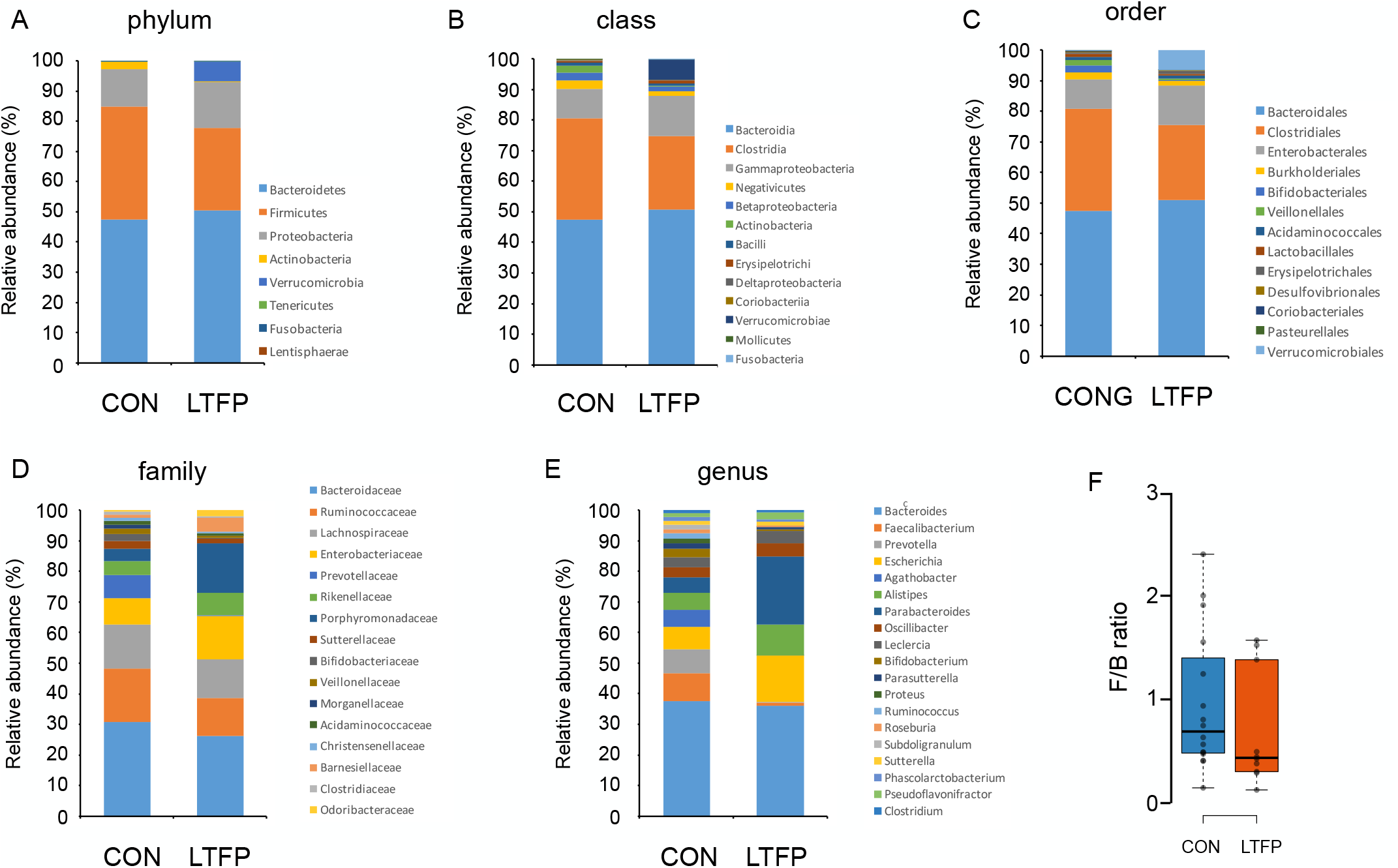
Relative abundance of gut microbiota in CON and LTFP group. The Stacked bar chart indicated the relative abundance of bacterial taxa in the gut microbiota of CON and LTFP groups based on 16S rRNA gene sequencing. Bacterial composition was shown at five taxonomic levels: (A) phylum, (B) class, (C) order, (D) family, and (E) genus. (F) *Firmicutes* / *Bacteroidetes* (F/B) ratio in CON and LTFP group.

Notably, *Firmicutes* (at the phylum level, p = 0.0424), *Ruminococcaceae* (at the family level, p < 0.0001), *Prevotellaceae* (at the family level, p = 0.0029), *Veillonellaceae* (at the family level, p = 0.0017), Roseburia (at the genus level, p = 0.0017), and *Subdoligranulum* (at the genus level, p = 0.0001) were significantly reduced in the LTFP group compared with those in the CON group; whereas *Ruminococcus* (at the genus level, p = 0.0143) and *Ruthenibacterium* (at an the genus level, p = 0.0048) were more abundant in the LTFP group (Table 2).

### Potential functional pathways associated with LTFP

We explored microbiota functions based on the metagenomes inferred by the PICRUSt algorithm. A principal component analysis based on 16S rRNA sequence-derived functional profiles was conducted to compare the microbiota of the CON and LTFP groups. In this analysis, the PC1 explained 38.2% of the variance, whereas the PC2 and PC3 explained 22.8% and 7.4% of such variance, respectively. As shown in Figure 4A, the CON and LTFP groups exhibited distinct clustering patterns along the PC1 axis. We compared the differences between the CON and LTFP groups and identified 63 MetaCyc pathways that showed significant differences (p < 0.05; Fig. 4, Table S3). Moreover, 27 pathways were found to be significantly different at p < 0.01 (Fig. 4, Table S3). In the LTFP group, the top Five enriched pathways were guanosine nucleotides degradation III, purine nucleotides degradation II, adenosine nucleotides degradation II, the urea cycle, and formaldehyde oxidation I. In contrast, in the CON group, the top Five enriched pathways were the superpathway of polyamine biosynthesis II; S-adenosyl-L-methionine cycle I; peptidoglycan biosynthesis IV; myo-, chiro-, and scillo-inositol degradation; O-antigen building blocks biosynthesis; and the superpathway of fucose and rhamnose degradation.

**Figure 4.**
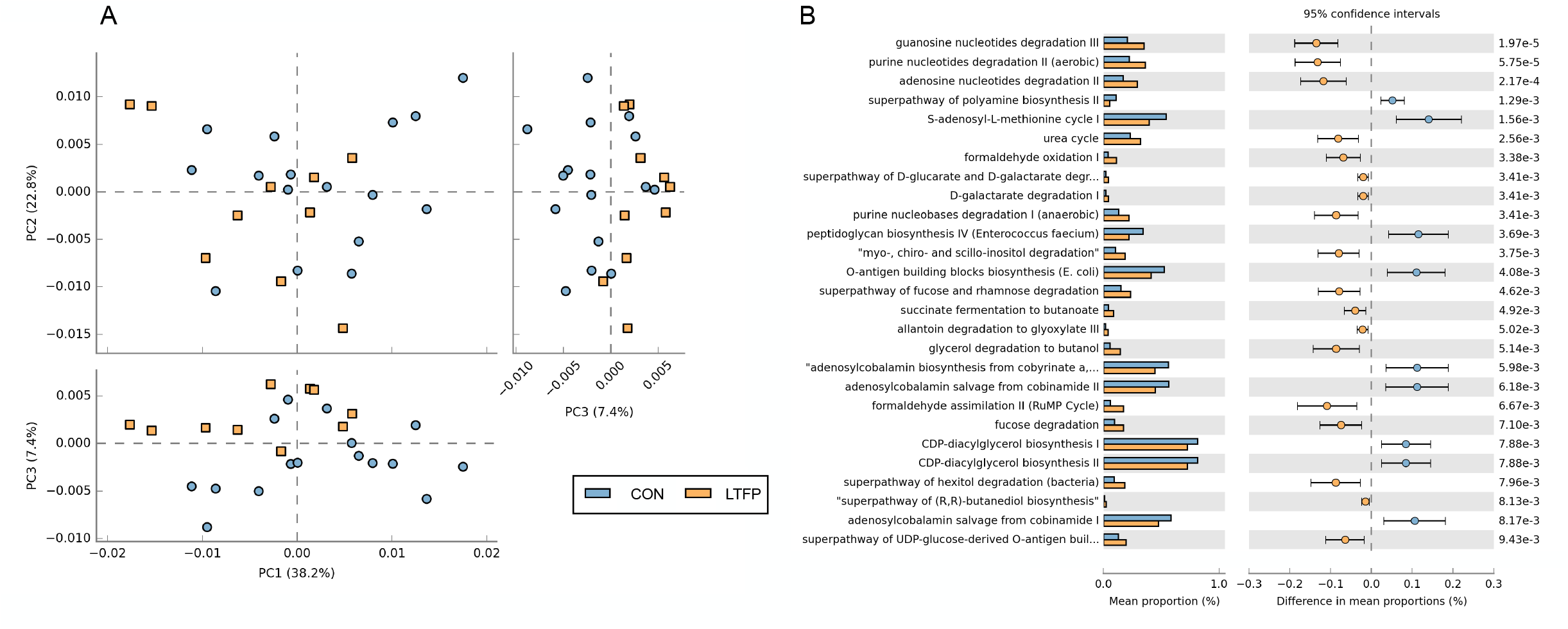
Functional predictions for the gut microbiota in the CON and LTFP group. (A)The PCoA of gut microbiota functional profiles based on 16S rRNA sequencing data. Blue circles represent the CON group (n=16), and orange squares represent the LTFP group (n=10). (B) The functional analysis was predicted from 16S rRNA gene-based microbial compositions. Statistical significance was determined using Welch’s t-test, with P < 0.05 considered significant.

## Discussion

In this study, we characterized the gut microbiota profiles of elderly Japanese patients undergoing LTF using 16S rRNA gene sequencing. Our results demonstrated that the microbial community in the fecal specimens of LTFP was significantly different from that of the CON group. Specifically, compared with that of the CON group, bacterial richness significantly decreased in LTFP. Moreover, the abundances of two genera increased, whereas those of one phylum, one order, three families, and three genera decreased in the LTFP group. Functional analysis revealed that nucleotide degradation pathways such as guanosine nucleotides degradation III and purine nucleotides degradation II, along with the urea cycle and formaldehyde oxidation, were enriched in LTFP.

### Decreased gut microbiota diversity

Previous studies have reported that TF decreases the diversity of gut microbiota (Gerasimidis et al., 2014; Quince et al., 2015). This reduction has been observed in both postoperative patients with gastric cancer and in patients with Crohn’s disease 2–4 weeks after TF. Our findings align with this trend, showing a decreased diversity in LTFP. The duration of TF and the specific dietary content of TF products, including fiber and oligosaccharides, are likely contributors to these changes (Yoshikawa, 1985; Wierdsma et al., 2009). However, we could not determine whether the decrease in diversity was a persistent effect or an adaptation to initial improvements in the disease state because early-phase samples were not obtained. Although a decrease in diversity is generally viewed negatively, some studies suggest that it may coincide with disease remission, complicating the interpretation.

### Changes in individual gut bacterial composition and characteristics of the elderly

The *Firmicutes*/*Bacteroidetes* (F/B) ratio has been proposed as a marker of the gastrointestinal microbiome status. In healthy adults, this ratio is approximately 10.9, but decreases to approximately 0.6 in the elderly (Mariat et al., 2009). In the present study, the median F/B ratio in healthy CON group members was 0.69, whereas that in LTFP was 0.44, with no significant difference observed. This finding suggests that LTF does not influence the F/B ratio in elderly individuals.

At the genus level, *Faecalibacterium* abundance was reduced in LTFP. As a known butyrate producer associated with mucosal protection, this decrease may contribute to gastrointestinal issues such as constipation and diarrhea. Contrary to previous reports, *Ruminococcaceae* and *Veillonellaceae* also decreased in patients from the LTFP group. Differences in age, ethnicity, and TF formulation may also account for these discrepancies.

### Changes in bacterial composition in elderly Japanese patients

The composition of the gut microbiota differs between populations from different geographical regions and ethnicities. Studies have shown that Japanese and other Asian populations possess distinct microbial profiles from Western populations (Abdill et al., 2025). Age-related shifts in the microbiota of Japanese individuals include increases in the abundance of *Proteobacteria* and decreases in that of *Firmicutes* (Odamaki et al., 2016). Our findings revealed a significant decrease in *Firmicutes* in LTFP, whereas *Proteobacteria* levels remained unchanged, suggesting that the changes were likely due to TF rather than aging. As *Firmicutes* include key short-chain fatty acid producers, such as *Ruminococcaceae* and *Roseburia*, their reduction may impair mucosal function. Similar patterns have been observed in chronic constipation and chemotherapy-induced diarrhea (Yu et al., 2023; Kawasaki et al., 2023). This overlap highlights a possible gut-microbiota-host interaction, although causality remains unclear.

### Functional alterations of gut microbiota in LTFP

Functional predictions based on inferred metagenomic profiles revealed an enrichment of catabolic pathways in LTFP. These included nucleotide degradation and fermentation pathways (e.g., hexitol degradation and glycerol fermentation), suggesting that the microbiota adapted to a fiber-limited environment. Conversely, the CON group showed an enrichment of biosynthetic pathways, including those involved in peptidoglycan and phospholipid synthesis as well as in amino acid and carbohydrate metabolism. These features are indicative of a stable, homeostatic gut environment supported by a fiber-rich diet.

## Limitations

Importantly, these findings cannot be extrapolated to all human groups. Other limitations of this study are as follows: (1) it was conducted on individuals from a specific age and ethnicity group, and the sample size was small; (2) no detailed study of the diet followed by CON group members was available; (3) the reasons for TF as well as the medical conditions and situations of the patients were not uniform; (4) the study design was cross-sectional and did not include baseline microbiota data from before the initiation of TF; and (5) the functional analysis was based on predicted metagenomic data, which may not accurately reflect the actual metabolic activity of the gut microbiota. Future studies with larger cohorts, longitudinal designs, detailed nutritional and medication records, and multi-omics approaches are warranted to better elucidate the complex interactions between LTF, gut microbiota, and gastrointestinal functions in elderly patients.

## Data Availability

All data produced in the present study are available upon reasonable request to the authors

## Acknowledgements

This work was supported by JSPS KAKENHI Grant Number JP 21K17333, JP 23K10588 and the Fundamental Research Funds form the Nayoro city university for YM, HT and YH. We would like to acknowledge Dr. Kazuhiro Sako and Dr. Yoko Nomura for critiques for our experiment. We would like to thank Editage (www.editage.jp) for English language editing.

## Author contributions

YM, MS, HT and YH designed the research. YM, MS, YO, YF and YH performed the experiments. YM, HT and YH wrote the article.

## Figure and reagent

***Table.S1.* Summary of 16S rRNA Sequencing and Alpha Diversity Metrics for Each Subject.** 16S rRNA sequencing data for CON and LTFP samples. This table summarizes sequencing results for each sample, including total reads, quality scores (Q20 and Q30), number of non-chimeric reads, observed OTUs, and Chao1 richness index.

***Table.S2. Taxonomic profile of gut microbiota in all samples***

***Table.S3. Predicted functional pathway abundances based on 16S rRNA gene sequences***

